# Studying Veteran food insecurity longitudinally using electronic health record data and natural language processing

**DOI:** 10.1101/2024.08.30.24312861

**Authors:** Alec B. Chapman, Talia Panadero, Rachel Dalrymple, Alicia Cohen, Nipa Kamdar, Farhana Pethani, Andrea Kalvesmaki, Richard E. Nelson, Jorie Butler

## Abstract

Food insecurity is an important social risk factor that is directly linked to patient health and well-being. The Department of Veterans Affairs (VA) aims to identify and resolve food insecurity through social and clinical interventions. However, evaluating the impact of such interventions is made challenging by the lack of follow-up data on Veteran food insecurity status. One potential solution is to leverage documentation of food insecurity in electronic health records (EHRs). In this paper, we developed and validated a natural language processing system to identify food insecurity status from clinical notes and applied it to study longitudinal trajectories of food insecurity among a large cohort of food insecure Veterans. Our analyses provide insight into the timing and persistence of Veteran food insecurity; in the future, our methods will be used to evaluate food insecurity interventions and evaluate VA policy.

## Introduction

Food insecurity, defined as a lack of access to affordable, healthy food,^1^ was reported by over 10% of US households in 2022.^2^ Individuals with food insecurity are at an increased risk for physical conditions such as diabetes and obesity^3–5^ and poor mental health outcomes.^6^ Recent work suggests that Veterans are more likely to live in a food insecure household than non-Veterans after adjusting for household and individual differences.^7^ The Department of Veterans Affairs (VA) has taken steps to address food insecurity among Veterans, including forming a central Food Security Office;^8^ implementing a universal screening program;^9,10^ establishing local food pantries on VA campuses;^11^ and developing public-private partnerships offering services to food insecure Veterans.^12,13^

Successfully addressing Veteran food insecurity requires measuring the prevalence across the U.S. Veteran population and evaluating the impact of interventions. However, there are challenges in measuring long-term food insecurity using traditional methods such as surveys and interviews, which can be expensive and time consuming. An alternative approach is to leverage data in the electronic health record (EHR). The VA EHR is a centralized repository for data collected from VA facilities across the nation, making it a valuable resource for conducting large-scale studies among the Veteran population. A key data element used to document food insecurity recorded in the VA EHR is the Food Insecurity Clinical Reminder, a screener administered at point of care with the intention of facilitating patient-level interventions and systematic measurement of food insecurity.^9,10^ Implementation of the screener has greatly improved the VA’s capability to monitor food insecurity in the Veteran population, as demonstrated by studies estimating the overall prevalence of food insecurity in VA and characteristics associated with food insecurity.^9,14,15^ However, the positivity rates of the food insecurity screener have been found to be lower than prevalence estimates from other studies and to vary substantially between VA medical facilities.^14^ Additionally, as the primary purpose of the screener is identifying cases of food insecurity and flagging them for clinical intervention, it is only administered every 6-12 months, which may limit its utility for measuring the persistence or resolution of food insecurity over time.

Assessment of Veteran food insecurity may be improved by leveraging free-text clinical notes. The response to a positive food insecurity screen involves an interdisciplinary team that includes primary care physicians, dietitians, social workers, and nurses. Food insecurity status and subsequent treatment decisions may be documented in clinical notes and can be extracted using natural language processing (NLP). Numerous studies have demonstrated the capability of NLP to extract social determinants of health (SDoH) from the EHR.^16–18^ In VA, NLP has been used to overcome limitations of structured EHR data when evaluating the effect of VA homeless programs on longitudinal housing status.^19–21^ However, to date, no studies have applied this approach to study food insecurity.

This paper describes an analysis of longitudinal food insecurity in the VA using a combination of structured and unstructured EHR data. We first developed an NLP system for extracting longitudinal food insecurity status from clinical notes and validated the system using reference standard data. This system was then applied to extract food insecurity status for up to two years following an initial positive screen. We then used this longitudinal dataset to examine the probability of being food insecure over the two-year follow-up period.

## Methods

Source code and supplemental materials are available on GitHub.^1^

### Data and Setting

We performed our analyses using data from the VA Corporate Data Warehouse (CDW). Our cohort comprised Veterans 18 years and older who had screened positive using the VA Food Insecurity Screener Clinical Reminder. The food insecurity screener was first implemented nationally in 2017 using a single-item question^9^ and updated to a validated 2-item screener in April 2021. ^22^ The food insecurity screener is meant to be performed at least annually for all non-institutionalized Veterans seen in primary care, although it may be administered in any VA clinical setting. Following a positive screen, Veterans are offered referrals to a social worker and/or dietitian, and staff are prompted to perform a follow-up screen 3 months later. If a Veteran screens positive for food insecurity again at this follow up screen, they are then due to be rescreened every 3 months for as long as they continue to screen positive. While these screening procedures are recommended for all national VA facilities, the degree to which these recommendations are followed is not well-studied.

We restricted our analyses to Veterans who had their incident positive screen between April 1^st^, 2021, and December 1^st^, 2021. For each Veteran who screened positive during this timeframe, we retrieved clinical data including co-morbidities and vitals in the 12 months prior to their incident screen, as well as follow-up food insecurity screens through 2 years after. This study was approved by the University of Utah Institutional Review Board.

### Note selection

We identified clinical notes containing keywords related to food insecurity that were authored between 30 days prior to an initial screen and up to two years after. The complete list of keywords is displayed in the **Supplemental Materials** (**Exhibit S1)**. For NLP development and validation, we excluded notes from dates that the Veteran had a food insecurity screen, as structured screening data were available on those days and templated text documenting a positive screen was populated in the note, essentially ensuring a positive classification. We identified whether notes were authored by any of three provider types who were identified in the literature as playing a critical role in responding to Veteran food insecurity,^9^ namely primary care physicians, dietitians, or social workers, and calculated descriptive statistics characterizing the distribution of authoring provider types.

### Annotation

Two authors (ABC and TP) iteratively developed an annotation schema for classifying whether notes contained documentation of food insecurity. Annotators met and reviewed a sample of texts to develop a classification scheme with three classes: *Positive, Negative*, or *Unknown*. This logic is displayed visually in **Exhibit S2** in the **Supplemental Materials** and described in detail below.

A document was classified as *Positive* if either:

1. The note explicitly states the Veteran is food insecure.
2. The note does not explicitly state the Veteran is food secure AND at least one of the following criteria are met:
  a. The Veteran is receiving food from a short-term assistance/emergency food relief program.
  b. The Veteran is applying for a long-term food assistance program.
  c. The Veteran has restricted food intake, is worried about future food intake, or eats low-quality food due to socioeconomic challenges.

The second part of this definition includes two types of food assistance. *Emergency food relief services* included food pantries, soup kitchens, food banks, or canned food drives. *Long-term food assistance programs* included federal programs such as the Supplemental Nutrition Assistance Program (SNAP, formerly the Food Stamp Program), Meals on Wheels, and the Special Supplemental Nutrition Program for Women, Infants, and Children. Connecting Veterans with these services is a key intervention for food insecurity;^9^ accordingly, enrollment in any of these long-term food assistance programs was not necessarily considered evidence of current food insecurity. However, if a note indicated a Veteran was applying to such a program, expressed a need for participating, or was still experiencing food insecurity despite receiving these benefits, then the note would be annotated as *Positive*.

A *Negative* classification was assigned if the criteria for *Positive* was not met and there was at least one of the following:

1. An explicit negation of food insecurity.
2. An explicit statement of food security.
3. A statement saying the Veteran had sufficient quantity or access to food.

If none of these criteria were met, the note was annotated as *Unknown*. The two annotators developed the guidelines in batches of 25-50 notes. After each batch, inter-annotator agreement (IAA) was measured using Cohen’s Kappa until a sufficient level of agreement (>0.8) was met for at least two iterations. After achieving sufficient IAA, 200 notes were annotated by one author (TP) and set aside for testing the NLP system.

### NLP system

We developed a rule-based NLP system, referred to as Food Insecurity in Electronic and Social Work Texts (FIIESCT) using the medspaCy Python library.^23^

#### Entity extraction

medspaCy’s *TargetMatcher* component was used to identify terms related to food insecurity including direct mentions of food insecurity or food security; long-term food assistance; emergency food relief; and access to food. Examples are shown in **Table 1**.

**Table 1.**
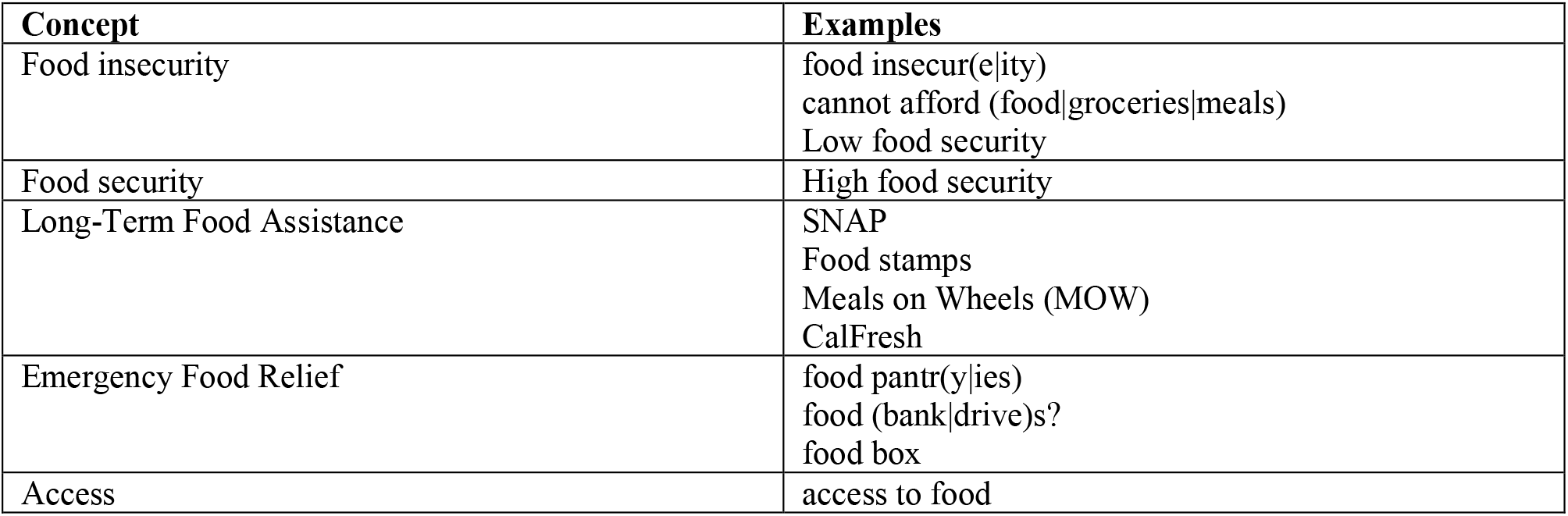
Semantic concepts and patterns including regular expressions extracted by FIIESCT.

#### Attribute detection

After extracting text spans representing food-related concepts, linguistic attributes were identified for each entity using the *ConText* algorithm^24^ and by identifying an entity’s location in the note using medspaCy’s *Sectionizer* component. The attribute types assigned to each entity are summarized in **Table 2** and included negation, temporality, and whether the mention referred to a hypothetical scenario. In addition, we defined two custom attributes for this project: first, whether a mention was expressed as a *need* (e.g., “he needs help with food”); and second, whether the text expressed the Veteran was *seeking* food assistance (e.g., “applying for SNAP benefits”). If none of these attributes were present, the entity was determined to be *asserted*.

**Table 2.** Linguistic attributes extracted by *medspaCy*.

| Attribute | Examples |
| --- | --- |
| is_negated | *No access to food.*<br>He does *not* have any current issues with *food insecurity*. |
| is_hypothetical | He will reach out <u>if</u> he has any further *food insecurity needs*. |
| is_historical | <u>History of</u> homelessness and *food insecurity*. |
| is_need | He *needs* assistance with *affording meals*.<br>He <u>would benefit</u> from *food stamps*. |
| is_seeking (Food Assistance only) | She is *applying for SNAP benefits*.<br>Social worker drove the Veteran to the *food stamp office*. |
| is_asserted | He *is experiencing food insecurity*.<br>Does the Veteran have *access to food*? *Yes*. |

#### Document classification

The final processing step determined a document-level classification of food insecurity status. A note was classified as *Positive* if there was at least one asserted mention of food insecurity, a negated mention of food security, an asserted mention of emergency food relief, or a mention of long-term food assistance with a *seeking* or *need* modifier. A note was classified as *Negative* if none of these concepts were present and there was either (1) a negated mention of food insecurity; or (2) an asserted mention of food security. If none of these criteria were met, the document was classified as “Unknown” and considered to be uninformative regarding their current food insecurity status.

#### Evaluation

We performed a final evaluation on the testing set of 200 notes. The author responsible for developing FIIESCT (ABC) was blinded to the testing set until the final evaluation. Performance measured using positive predictive value (PPV), sensitivity, and F1-score within each of the three document classes with bootstrapped 95% confidence intervals for each statistic. Additionally, we performed an error analysis by reviewing misclassified documents in the testing set and determining the primary cause of misclassification.

### Longitudinal analysis

After validating the NLP system, we applied FIIESCT to model the risk of experiencing food insecurity each day after an initial positive screen over two years. We used an approach that has previously been used to study longitudinal housing outcomes in VA. ^19–21^ This approach consisted of three steps: first, extracting a Veteran’s food insecurity status using both the food insecurity screener and FIIESCT; second, modeling the visit process and calculating inverse intensity weights to account for the irregularity of food insecurity documentation; and third, modeling the overall probability of food insecurity among the entire cohort as a smooth function of time.

#### 1. Extracting visit-level food insecurity

For each Veteran in the cohort, we retrieved all documents containing keywords related to food insecurity as described above. Each note was assigned a document classification, and notes classified as *Unknown* were excluded from further analyses. We defined food insecurity-related visits as any visit on which the Veteran had a food insecurity screener result or at least one note classified by FIIESCT as *Positive* or *Negative*. Visits were classified as positive if either the screener was positive or at least half of the notes included on that date were classified as *Positive*.

#### 2. Calculating inverse intensity weights

A Veteran’s food insecurity status in the EHR at irregular time intervals. Furthermore, we hypothesized assessments of food insecurity would occur more frequently for Veterans who are currently food insecure, as they would be more likely to be receiving services or discussing food insecurity with their providers. This imbalance in assessment times, referred to as informative assessment times, can lead to biased estimates of a longitudinal outcome variable. To address this issue, we incorporated inverse intensity weighting (IIW) into our analysis.^25^ Briefly, IIW assumes that the frequency with which a Veteran visits and has documentation of food insecurity is independent of their current food insecurity status conditional on observed covariates and food insecurity history. The visit process is modeled using a recurrent event model and used to estimate weights proportional to the inverse of the intensity (i.e., instantaneous probability) of visiting at each observed time point. Variables in the recurrent event model included Veteran demographics, co-morbidities, vitals, and the Veteran’s last recorded food insecurity status.

#### 3. Modeling the probability of food insecurity over time

We next fit two logistic regression models to estimate the probability of being food insecure each day during the two years after an initial screen. Food insecurity status at each food insecurity-related visit was modeled as a function of time using cubic splines with the model weighted by inverse intensity weights. Bootstrapping was used to compute 95% confidence bands. The first model included both screener results and clinical notes, while the second model used screener results alone. We assessed the impact of supplementing the screener with NLP by comparing the estimated probabilities from the two models over the two-year period.

## Results

### Cohort

Our cohort consisted of 29,525 Veterans. Veteran characteristics are displayed in **Table 3**. Veterans were primarily white and male, and most were older than 40. There was a relatively high prevalence of homelessness and mental health diagnoses, as well as clinical co-morbidities including hypertension and hyperlipidemia.

**Table 3.** Characteristics of Veterans who screened positive for food insecurity.

| Characteristic | N = 29,525<sup>1</sup> |
| --- | --- |
| Gender |  |
| Male | 25,101 (85%) |
| Female | 4,424 (15%) |
| Age |  |
| <40 | 7,068 (24%) |
| 40 to 49 | 4,105 (14%) |
| 50 to 59 | 5,774 (20%) |
| 60+ | 12,578 (43%) |
| Race |  |
| White | 16,367 (55%) |
| Black/African American | 9,878 (33%) |
| American Indian/Alaska Native | 499 (1.7%) |
| Native Hawaiian/Other Pacific Islander | 448 (1.5%) |
| Asian | 374 (1.3%) |
| Missing | 1,959 (6.6%) |
| Ethnicity |  |
| Not Hispanic or Latino | 25,587 (87%) |
| Hispanic or Latino | 2,514 (8.5%) |
| Missing | 1,424 (4.8%) |
| BMI |  |
| <18.5 | 472 (1.6%) |
| 18.5 to 24.9 | 5,898 (20%) |
| 25 to 29.9 | 7,212 (24%) |
| 30 to 39.9 | 7,769 (26%) |
| 40+ | 1,621 (5.5%) |
| Missing | 6,553 (22%) |
| Medical History |  |
| Hypertension | 8,986 (30%) |
| Hyperlipidemia | 7,240 (25%) |
| Type II Diabetes | 5,252 (18%) |
| History of homelessness | 10,883 (37%) |
| Substance use disorder | 7,343 (25%) |
| Any mental health diagnosis | 14,577 (49%) |

### Annotation

The final IAA measured during development was Kappa=0.865, indicating high agreement between the two annotators. The final testing set of 200 notes contained 80 positive documents, 36 negative documents, and 88 unknown documents. The relatively high number of unknown labels were included in the annotation set due to either templated texts containing food insecurity/food assistance-related keywords (e.g., program intake forms with a spot for recording food stamp benefits) or ambiguous terms (e.g., the abbreviation “SNAP” referring to patient “Strengths, Needs, and Preferences”).

### Document classification

The PPV, sensitivity, and F1 for each document class are shown in **Table 4**. FIIESCT achieved moderate performance in each category. The lowest-performing category was *Negative*, with both PPV and sensitivity of 0.686. *Positive* documents had higher PPV (0.787) and sensitivity (0.747). *Unknown* documents had the highest values in both metrics. An analysis of errors in the testing set is included in **Exhibit S3** of the **Supplemental Materials** with the most common sources of errors being incorrect attribute detection by ConText, reference standard labels that were inconsistent with the annotation guidelines, and food insecurity-related terms not included in the NLP lexicon.

**Table 4.** Document classification performance metrics and 90% confidence intervals using 200 testing documents.

|  | PPV | Sensitivity | F1 | Count |
| --- | --- | --- | --- | --- |
| **Positive** | 0.787<br>(0.730, 0.844) | 0.747<br>(0.683, 0.811) | 0.766<br>(0.719, 0.814) | 79 |
| **Negative** | 0.686<br>(0.588, 0.784) | 0.686<br>(0.589, 0.782) | 0.686<br>(0.607, 0.764) | 35 |
| **Unknown** | 0.800<br>(0.745, 0.855) | 0.837<br>(0.786, 0.888) | 0.818<br>(0.777, 0.860) | 86 |
|  |  |  |  | 200 |

### Longitudinal analysis

#### Food insecurity-related visits and classifications

We next extracted food insecurity status for the entire Veteran cohort and examined food insecurity-related visits and classifications longitudinally. Descriptive figures of the longitudinal dataset are shown in **Figure 1**.

**Figure 1.**
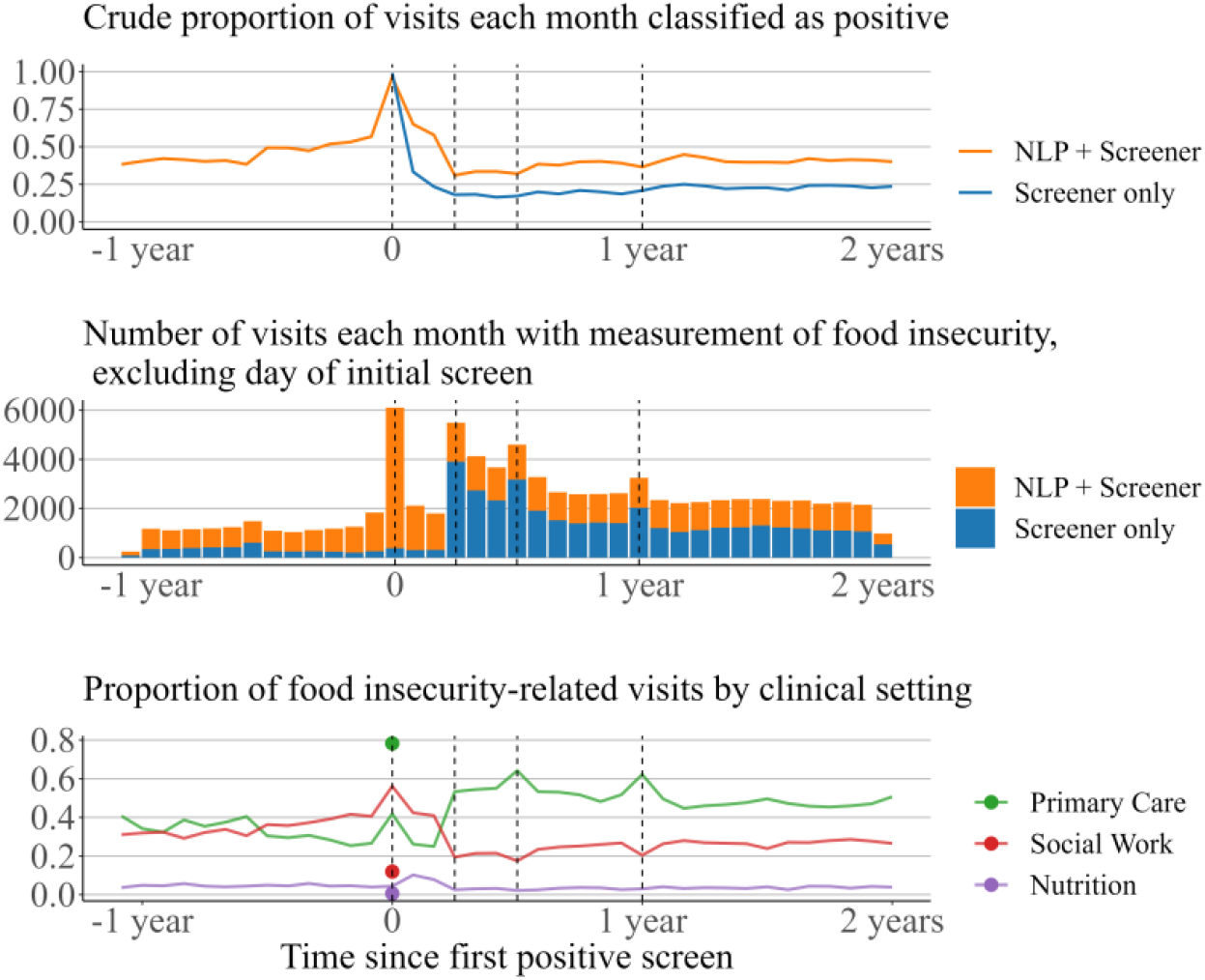
Descriptive figures of food-insecurity related visits between 1 year prior and up to 2 years after an incident positive screen, with vertical lines showing the time of the first screen and times of scheduled follow-ups at 90 days, 180 days, and 365 days.

Among the 29,525 Veterans in our cohort, there were 71,336 food insecurity-related visits in the two years following their incident positive screen and 15,180 in the year prior. Beginning 30 days before the positive screen, the NLP showed an increase in the number of food insecurity-related visits and the proportion classified as positive. During the 90 days after the incident screen, the frequency of notes decreased, as did the positivity rate. Screens were infrequent compared to clinical notes and showed a lower positivity rate than NLP throughout follow-up. Visits during which a screener was administered showed high agreement between the screener and NLP visit classifications (98.4%) **(Supplemental Materials Exhibit S4)**. Counts of both NLP and screenings increased at scheduled follow-up times (90, 180, and 365 days). Most initial screens were performed during visits with a primary care physician. During the preceding 6 months and the following 90 days, most food insecurity-related visits involved an encounter with social work, while primary care was the most common setting after 90 days. Nutrition visits were rare but increased slightly during the 90 days after an initial screen.

#### Model results

**Figure 2** displays the estimated probabilities of food insecurity obtained using inverse intensity-weighted models with NLP and screener-derived outcome variables. The estimated coefficients and 95% confidence intervals for the recurrent event model are displayed in **Exhibit S4** in the **Supplemental Materials**. When including NLP, the probability of food insecurity decreased sharply during the first 30 days (estimated probability on day 30 0.761; 95% confidence band [0.733, 0.788]) and continued to decrease through day 90 (0.330; [0.301, 0.354]). The estimated risk continued to decrease until reaching the lowest value at day 139 (0.317; [0.289, 0.344]). There was then a slight increase during the final 1.5 years (estimated probability at 2 years of 0.339; [0.312, 0.367]).

Using only screener data resulted in lower estimates of food insecurity. The screener showed a probability at day 30 of 0.391 [0.312, 0.464] and at day 90 of 0.141 [0.068, 0.214]. There was then an increase in probability following the 90-day follow up time, with an estimated probability on day 183 of 0.196; [0.123, 0.269]. Like the NLP, the screener showed a steady risk of food insecurity during the final year of the study, with a final estimated probability of 0.185 [0.112, 0.258]. Due to the lower frequency of screening compared to documentation in clinical notes, the model using screener data only had less precise estimates (i.e., larger 95% confidence bands) than the model using NLP.

**Figure 2.**
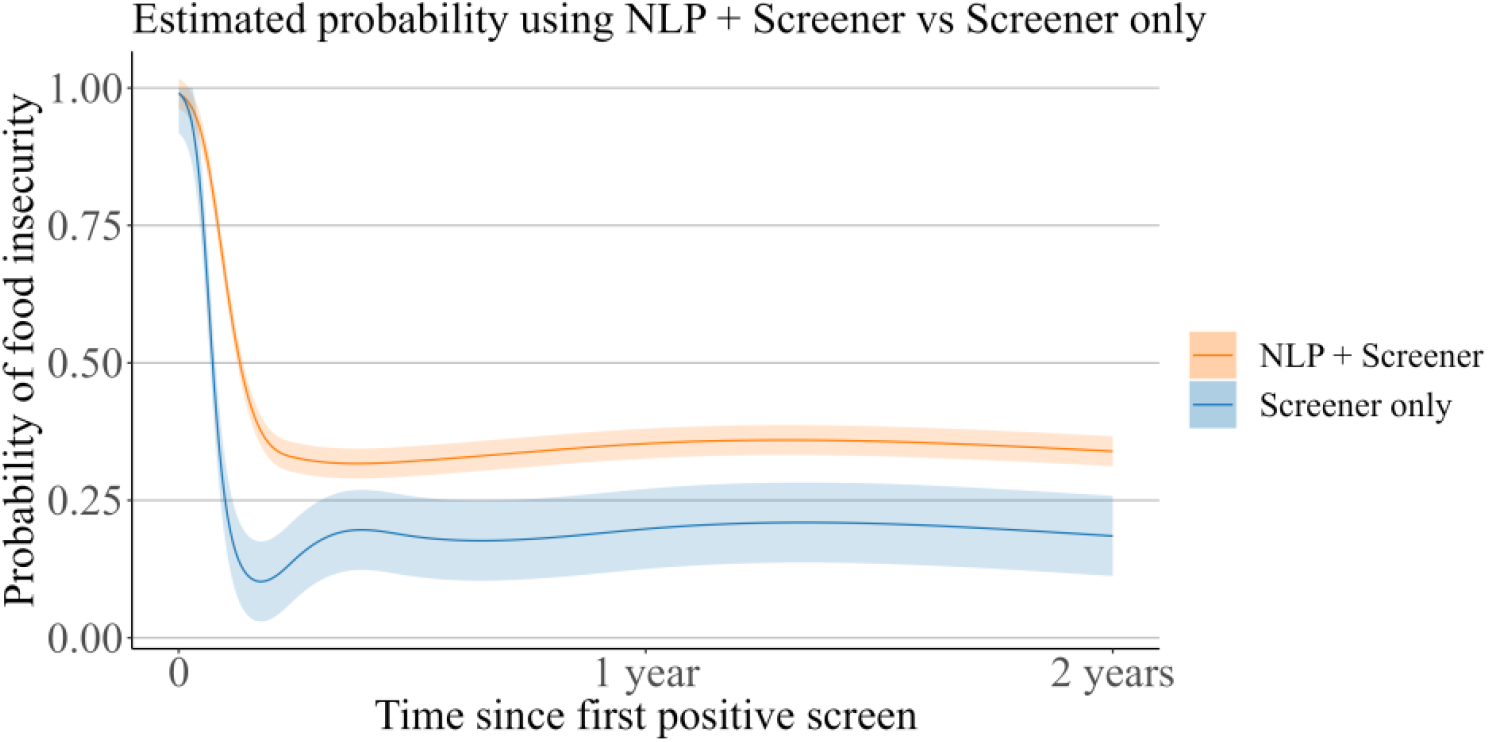
Estimated probability of food insecurity over the two years after an initial positive screen.

## Discussion and Conclusions

### Discussion

In this study we developed FIIESCT, an NLP system for identifying food insecurity status from clinical notes. We then applied FIIESCT to construct a longitudinal dataset combining information in clinical notes with structured data and examined long-term food insecurity for a cohort of recently food insecure Veterans. Compared to using screening data alone, FIIESCT showed higher estimates of food insecurity over two years following a positive screen. This has important implications for VA policymakers and researchers. Nearly one third of Veterans in our sample were estimated to be experiencing food insecurity two years after being identified as food insecure.

Addressing this sustained risk may require increasing outreach to Veterans after they screen positive to ensure they are connected with services needed for long-term food security, such as nutrition education or produce prescription programs.^26^

This study also demonstrates the value of using NLP to study food insecurity longitudinally. The screener is the standard method for measuring food insecurity in VA; however, our findings suggest limitations of screener data. Food insecurity was recorded more frequently in clinical notes, particularly before the Veteran’s first positive screen and between recommended follow up times. Clinical notes also contained more frequent positive documentation of food insecurity throughout the study period, with NLP showing a nearly two-fold increase in risk compared to the screener two years after an initial positive screen.

Several factors may contribute to the differences in the frequency and positivity of clinical notes and the screener. First, the screener was designed to be administered periodically at pre-specified follow-up times, while clinical notes are typically documented at any visit and may include information relevant to food insecurity status. This is demonstrated by the large increase in food insecurity assessments around the 90-day follow up time: very few screens were performed between the initial screen and 90-day follow-up and clinical notes were documented far more frequently. The interval the positive screen and 90-day follow-up is a critical time during which Veterans are connected with VA and community services,^9^ making this a crucial time to assess Veterans’ engagement and access to services. Furthermore, the increase in assessments after 90 days was accompanied by a sharp increase in the positivity rate for the screener. A possible explanation of this trend is that the few screening results performed documented before the intended follow-up time may not be representative of the food insecurity status for the larger cohort, even after applying inverse intensity weights. In contrast, this trend was not observed in the NLP-derived estimates, suggesting that information in clinical notes may provide more reliable estimates of food insecurity during this important time period.

Second, the two methods of measuring food insecurity may differ due to variation in documentation practices between different types of providers. While the screener may be administered by any VA provider, it is only mandated during primary care visits. However, food insecurity is a complex phenomenon that requires coordinated responses across clinical care teams.^9^ We found that clinical notes documenting food insecurity were frequently authored by social workers, particularly immediately before and an initial positive screen. Visits with dieticians increased during this time but were rare throughout the entire study period. While Veterans who screen positive are prompted to consult with a dietitian, these findings suggest that few Veterans do so. Future work should more closely examine the process by which VA providers respond to Veteran food insecurity and consider challenges that providers and Veterans face in responding to food insecurity.

Third, the definition of NLP used when designing the NLP differs somewhat from the definition used in the screener. Our definition incorporated information about food assistance services and barriers to food access in addition to explicit diagnoses of food insecurity. In contrast, the criteria for a positive food insecurity screen requires the Veteran endorsing a particular experience of food insecurity – specifically, being worried that their food would run out and not having money to buy more. While Veterans regularly accessing a food pantry may not screen positive for food insecurity based on the above definition, that individual may still be of interest when identifying Veterans who would benefit from additional assistance. A future application of FIIESCT could be identifying Veterans with evidence of food insecurity in clinical notes who lack a positive food insecurity screen, allowing VA to better ascertain the prevalence of food insecurity, identify Veterans who need services, and study barriers in administering the screener to Veterans. Furthermore, the modular, transparent design of our NLP system facilitates implementing alternative definitions for different applications or to transport FIIESCT to other institutions while customizing for differing documentation practices and EHRs.

### Limitations

Our study had several limitations. First, our analysis was limited to the VA healthcare system. Findings may not generalize to other populations with different patient demographics or clinical practices. Second, our analysis did not account for measurement error in the NLP. FIIESCT achieved higher PPV and sensitivity for identifying *Positive* and *Unknown* documents but achieved lower performance for negative documents, which could account for some of the difference between the screener and NLP results. We focused on rule-based methods in this study with the intent of creating a flexible pipeline that could be modified in future applications; however, accuracy could be improved by leveraging statistical NLP models (e.g., large language models). Future work should consider other NLP methods and incorporate adjustments for measurement error in the outcome model.

### Conclusions

We developed a natural language processing system for extracting food insecurity from clinical notes and applied it to study long-term food insecurity outcomes following a positive food insecurity screen. FIIESCT identified recurring instances of food insecurity status not captured using structured data alone and led to higher estimates of long-term food insecurity. Future work will apply these methods to enhance screening of food insecurity in VA and to evaluate interventions targeting food insecurity.

## Data Availability

Due to the protected nature of health information, data is unavailable to the public. Aggregate data may be available upon request.

https://github.com/abchapman93/fiiesct_nlp

## Acknowledgements

This work was supported by the IDEAS Center and Veterans Affairs Health Services Research & Development. AJC was additionally supported by Grant Number CDA 20–037 from the Department of Veterans Affairs Health Services Research & Development Service. NPK was additionally supported by Grant Number CDA 21-032 from the Department of Veterans Affairs Health Services Research & Development Service.

## Footnotes

1 https://github.com/abchapman93/fiiesct_nlp

## Notes

### Competing Interest Statement

The authors have declared no competing interest.

### Author Declarations

Ethics committee/IRB of University of Utah gave ethical approval of this work.

### Summary of Updates

Slight edits and grammar fixes.

