## Supplemental Materials for "Studying Veteran food insecurity longitudinally using electronic health record data and natural language processing"

**Exhibit S1.** Keywords used for identifying documents related to food insecurity.

| | |
|---|---|
| food (in)?security | food access |
| food (in)?secure | supplemental nutrition assistance program |
| afford food | snap |
| sufficient food | food stamp |
| lack of food | food bank |
| limited food | food assistance |
| insufficient food | food resources |
| inadequate food | |

**Exhibit S2.** Document classification logic specified in the annotation guidelines.

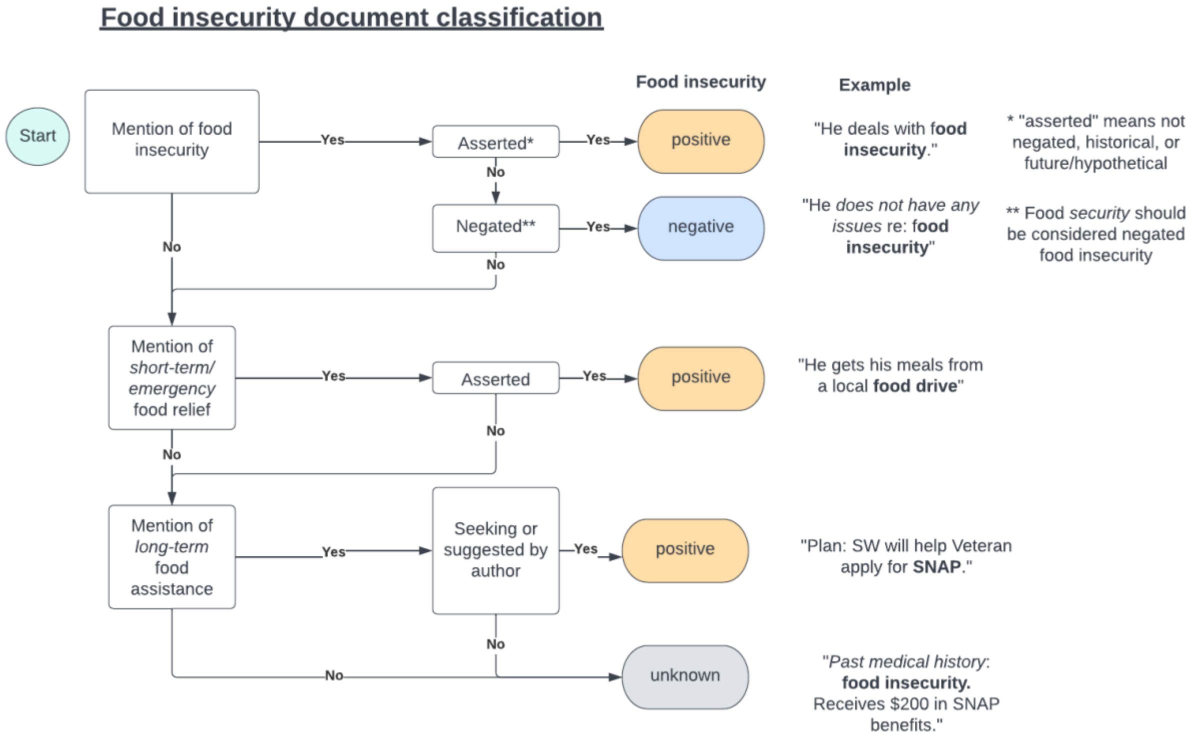


**Exhibit S3.** Error analysis in document classification testing set.

The most frequent cause of error was incorrect entity-modifier relationships assigned by *ConText*. For example, a note with the phrase "struggling with food security" was incorrectly classified as *Negative* instead of *Positive* because "struggling with" was not linked to food security. Similarly, the phrase "pending" was not captured by FIIESCT and was used to indicate a Veteran was in the process of applying for long-term food assistance, which would qualify the note as *Positive*. Some documents were correctly classified by FIIESCT but assigned an incorrect annotation. Although agreement was high between the two annotators (IAA = 0.865), the testing set was only annotated by a single annotator, increasing the possibility of annotator error.

Another common cause of error was missing or incorrect entities. This relates to the complexity of SDoH and social services offering support for food insecurity, as there was large variation in the way food insecurity was described or specific services that indicate food insecurity (e.g., names of specific food pantries or local emergency-relief programs). A common missed entity indicating *Negative* food insecurity was “adequate food”. This phrase had been deliberately excluded from the lexicon as it was the cause of false negative classifications in development notes belonging to food insecure Veterans. In these cases, “adequate food” referred to a having enough food for a short amount of time (e.g., for the rest of the week) rather than overall food security. Excluding this rule thus had a trade-off of causing false positive classifications in the testing set. Some incorrect entities were specifically related to semi-structured templates, such as forms that appeared to be copied and pasted into notes and mentioned food insecurity. Other causes of errors included non-explicit references to food insecurity that required additional interpretation or conflicting information in notes.

| **Error category** | **Description and examples** | **Count in testing set** |
|---|---|---|
| **Incorrect/missing modifier** | **A linguistic modifier was not identified or was identified and incorrectly linked to an entity.**<br><br>“Food stamp benefits are <u>pending</u>.”<br><br>“<u>struggling with</u> food security”<br><br>“<u>Seeking assistance</u> with stable housing and food resources”<br><br>“<u>Assistance needed</u> with food and housing”<br><br>“SW alerted by provider to Veteran’s <u>need for</u> food and housing resources” | 16 |
| **Incorrect annotation** | **Annotator’s document classification was inconsistent with guidelines due to annotator error or confusing text** | 13 |
| **Incorrect/missing entity** | **An entity was not identified or was incorrectly identified.**<br><br>“Veteran reported utilizing local resources such as [specific assistance facility name]”<br><br>“Veteran reported <u>adequate food</u>” | 6 |

| | | |
|---|---|---|
| | "Hasn't needed any additional food"<br><br>"food stamp appointment" ["appointment" indicates seeking services] | |
| **Template** | **Semi-structured forms or note headers that were not correctly parsed or ignored by NLP**<br><br>"Food insecurity: Getting $100 vouchers monthly; doesn't think food insecurity will be a concern"<br><br>"Food stamps: Yes<br>Food Pantry:<br>Transportation: Bus"<br><br>"Non-cash benefits/access to food - $150" | 5 |
| **Unclear text/non-explicit statement of food insecurity** | "Veteran reported she is unable to cook due to lack of space so she often eats fast food"<br><br>"SW called to inquire if patient needed assistance obtaining food. Patient disclosed he lost his debit card and was waiting for a new one."<br><br>"Writer asked if Veteran had sufficient food to last one week. Veteran reported he receives three meals a day." | 4 |
| **Classification logic** | **General logic for classification did not generalize to a particular document.**<br><br>"Food access/financial concerns: Yes.[food insecurity] Reports he has enough food[food security]" [false negative classification] | 1 |
| **Total** | | 45 |

**Exhibit S4.** Cross-tabulation of screener and NLP classifications during the two years following an initial positive screen.

| | | NLP | | | |
|---|---|---|---|---|---|
| Screener | | No visit | Negative | Positive | Total |
| No visit | Count | -- | 12,215 (12.5%) | 22,912 (23.4%) | 35,127 (35.9%) |

| Screener | | NLP | | | |
|---|---|---|---|---|---|
| | | No visit | Negative | Positive | Total |
| | Mar. pct [1] | -- | 30.1%; 34.8% | 40.5%; 65.2% | |
| Negative | Count | 62 (0.1%) | 28,183 (28.8%) | 491 (0.5%) | 28,736 (29.3%) |
| | Mar. pct | 7.0%; 0.2% | 69.5%; 98.1% | 0.9%; 1.7% | |
| Positive | Count | 822 (0.8%) | 162 (0.2%) | 33,120 (33.8%) | 34,104 (34.8%) |
| | Mar. pct | 93.0%; 2.4% | 0.4%; 0.5% | 58.6%; 97.1% | |
| Total | Count | 884 (0.9%) | 40,560 (41.4%) | 56,523 (57.7%) | 97,967 (100.0%) |

[1] Columns and rows percentages

**Exhibit S4.** Estimated intensity ratios (IRs) and 95% confidence intervals (CIs) for having a food insecurity-related visit.

| Characteristic | IR[1] | 95% CI[2] |
|---|---|---|
| Last visit classified as food insecure | 1.92 | 1.86, 1.99 |
| Gender | | |
| Male | -- | -- |
| Female | 1.08 | 1.04, 1.12 |
| Age | | |
| <40 y | -- | -- |
| 40 to <50 y | 1.15 | 1.09, 1.21 |
| 50 to <60y | 1.26 | 1.20, 1.32 |
| >=60 y | 1.35 | 1.29, 1.41 |
| Race | | |
| White | -- | -- |
| Black/African American | 1.04 | 1.01, 1.08 |
| American Indian/Alaska Native | 0.96 | 0.84, 1.10 |
| Native Hawaiian/Other Pacific Islander | 1.09 | 0.95, 1.26 |
| Asian | 0.96 | 0.85, 1.09 |
| Other/Missing | 1.02 | 0.96, 1.09 |
| BMI | | |
| 18.5 to 24.9 | -- | -- |
| <18.5 | 1.10 | 0.99, 1.24 |
| 25 to 29.9 | 0.94 | 0.90, 0.99 |
| 30 to 39.9 | 0.94 | 0.90, 0.99 |
| 40+ | 1.03 | 0.95, 1.11 |
| Missing | 0.97 | 0.93, 1.02 |
| Medical history | | |
| History of homelessness | 1.78 | 1.72, 1.84 |
| Hypertension | 1.09 | 1.05, 1.13 |
| Hyperlipidemia | 1.05 | 1.01, 1.09 |
| Type II Diabetes | 1.07 | 1.02, 1.12 |
| Substance use disorder | 1.10 | 1.06, 1.14 |
| Any mental health diagnosis | 1.11 | 1.07, 1.15 |

[1]IR=intensity ratio; [2]CI=confidence interval
